# Predictive Modeling of Resistant Hypertension Risk: Incorporating the TyG Index and Clinical Factors

**DOI:** 10.1101/2023.11.03.23298071

**Authors:** Hai-Tao Yang, Jing-Kun Liu, YI Yang, Ying-Ying Zheng, Xiang Xie

## Abstract

**Background:** Resistant hypertension (RH), a form of high blood pressure that remains uncontrolled despite maximum medication, poses a significant cardiovascular risk. This paper introduces a novel predictive model, combining the triglyceride-glucose (TyG) index with traditional clinical factors, to anticipate the development of RH in patients with newly diagnosed primary hypertension.

**Methods:** The study included hospitalized patients with newly diagnosed primary hypertension and stable blood pressure after medication treatment from August 2019 to early August 2021. After screening, a total of 1635 cases were finally included and divided into development and validation cohorts. The least absolute shrinkage and selection operator (LASSO) regression was applied to select potential risk factors. Multivariate Cox regression analysis was used to identify independent hazard factors constructed by the predictive nomogram. Receiver operating characteristic curve analysis (ROC), calibration curve, and C-index were used to evaluate the performance of the nomogram.

**Results:** A total of 1227 patients were assigned to the development queue, while 408 patients were assigned to the validation queue. The constructed column line chart includes five clinical variables: age, apnea-hypopnea index (AHI), uric acid, fasting blood glucose, and TyG index. Multivariate Cox regression analysis revealed that compared to the other four risk factors, TyG index (HR=3.97, 95% CI: 2.81 - 5.62, P < 0.01) was significantly associated with RH. ROC curve analysis showed prediction values of 0.895 and 0.837 for RH in the development cohort and prediction values of 0.854 and 0.832 in the validation cohort respectively. The C-index was found to be 0.76 in the development cohort and 0.66 in the validation cohort. Furthermore, Kaplan-Meier analysis indicated that compared to the low-risk group, there was a higher likelihood of developing RH in the high-risk group.

**Conclusions:** Based on the TyG index and electronic health record data, a model can be constructed to accurately and reliably predict the occurrence of RH in patients with stable blood pressure after initial diagnosis of primary hypertension and drug treatment.

## Introduction

Hypertension (HTN) is a pivotal modifiable risk factor and remains the leading cause of mortality [1,2]. Resistant hypertension (RH), a subtype of HTN, is characterized by the challenge of achieving blood pressure goals below 140/90 mmHg despite the maximum dose of three antihypertensive medications, including diuretics, or achieving blood pressure goals while on four antihypertensive medications [3]. RH affects approximately 12% to 15% of hypertensive individuals and carries a 47% to 200% higher risk of adverse cardiovascular events compared to non-RH [4]. Thus, early detection and timely intervention for RH are essential for reducing cardiovascular events.

As widely recognized, a causal association between insulin resistance (IR) and both HTN and cardiovascular events exists, potentially serving as a pivotal factor in the pathogenesis of HTN [6, 7]. In the clinical diagnosis domain, the progressive manifestation of RH among patients with HTN is observed, and it is regarded as an advanced stage of this disease state [5]. Further elucidation is required regarding the key factors contributing to the progression of RH. Currently, there is a dearth of clear reports that establish a causal relationship between IR and RH. Higher incidence of IR in patients with RH compared to control groups without RH has been observed in some studies [8]. However, other studies have reported that, despite significant reductions in blood pressure following renal denervation intervention for patients with RH, there was no improvement in IR [9]. Clarifying the causality between IR and RH holds significant potential for developing predictive mechanisms to identify high-risk hypertensive patients with RH and optimize their treatment while enhancing prognosis. This endeavor bears profound significance in terms of optimizing medical resources.

In recent years, numerous studies have demonstrated the effective quantification of IR levels and predictive value in cardiovascular disease prognosis by employing the triglyceride-glucose (TyG) index [10,11]. Given that TyG serves as an easily obtainable and reliable biomarker, and considering the necessity for a risk quantification model to determine RH based on real clinical data among hypertensive patients, we included patients with newly diagnosed HTN who exhibited stable blood pressure after long-term medication treatment for subsequent follow-up observation. Consequently, we investigated the combined predictive value of TyG and electronic health record (EHR) data concerning RH occurrence.

## Methods

### Study Design and Participants

Patients who were initially diagnosed with HTN at the Fifth Affiliated Hospital of Xinjiang Medical University from August 2019 to August 2021 and whose blood pressure remained stable after drug treatment were enrolled. The patients were admitted to the hospital for the exclusion of secondary hypertension. The inclusion criteria for the study were as follows: Biochemical tests and respiratory sleep tests were conducted during the initial hospital visit. Each adjustment of antihypertensive drug therapy was performed either on an outpatient or inpatient basis within our medical facility. Patients strictly adhered to prescribed medication for an extended duration, with no instances of noncompliance exceeding three consecutive days. Furthermore, they maintained regular sleep patterns without engaging in prolonged late-night activities and limited their average daily working time to less than 10 hours. Exclusion criteria included secondary HTN or white-coat HTN, coronary heart disease, diabetes mellitus, receiving noninvasive ventilation, abnormal liver and kidney function, cardiogenic shock, congestive heart failure, minor children, or pregnant women. A total of 1635 patients were ultimately enrolled in this retrospective cohort study and randomly divided into the Development cohort and Validation cohort at a ratio of 4:1 using a random number allocation method. These datasets were utilized to develop and validate a nomogram for predicting the development of treatment-RH. As this study was conducted retrospectively based on real-world situations, informed consent from the patients was not required. The study protocol was approved by the Ethics Committee of the Fifth Affiliated Hospital of Xinjiang Medical University (2022XE0103-1).

### Diagnosis of Hypertension

In the absence of antihypertensive medications, a diagnosis of HTN can be established if the systolic blood pressure is ≥140 mmHg and/or the diastolic blood pressure is ≥90 mmHg on three separate occasions [12]. To exclude white-coat HTN, dynamic blood pressure monitoring was conducted during the visit. The diagnostic criteria for HTN using dynamic blood pressure monitoring are as follows: 24-hour blood pressure ≥130/80 mmHg, daytime blood pressure ≥135/85 mmHg, or nighttime blood pressure ≥120/70 mmHg [13]. Primary HTN was diagnosed after ruling out secondary HTN through inpatient testing performed by an attending physician and a chief physician. A comprehensive treatment plan for HTN was collaboratively developed using angiotensin-converting enzyme inhibitors (ACEIs), angiotensin receptor blockers (ARBs), calcium channel blockers (CCBs), beta-blockers, and diuretics. Diuretics were added when a combination of three types of antihypertensive drugs was employed [14].

### Basic Information Collection, Plasma Biochemical Testing, and Respiratory Sleep Monitoring

The blood pressure measurements in this study pertain to the stable blood pressure levels of patients who have received antihypertensive medication. The blood pressure was assessed using an electronic sphygmomanometer by trained medical professionals, with readings taken in the morning after a minimum of 10 minutes of rest in a calm state for three consecutive days, and the mean value was calculated. Smoking was defined as continuous or cumulative tobacco use for six months or longer [15]. Furthermore, individuals consuming over 30 grams of alcohol per day were categorized as current drinkers. Plasma biochemical tests were conducted on fasting patients during their hospitalization period to evaluate results from blood routine examinations, liver and kidney function, lipid profile levels, and fasting blood glucose levels. Additionally, we calculated the TyG index using fasting triglyceride and fasting glucose levels according to the formula ln[TG(mg /dL) × FPG (mg/ dL)/2][16]. Polysomnography (Grael; Compumedics, Melbourne VIC Australia) was conducted for all study participants to meticulously record parameters, including airflow rate, thoraco-abdominal movements, pulse oximetry readings, and episodes of snoring. Subsequently, the apnea-hypopnea index (AHI) was calculated by considering the number of apneas and hypopneas per hour of sleep.

### Diagnosis of Resistant Hypertension

The study observed the endpoint event of RH, which was diagnosed by a attending physician and a chief physician through two or more outpatient visits. In addition to lifestyle improvements, a combination of three different types of antihypertensive drugs is usually used, including a long-acting calcium channel blocker (CCB), an angiotensin-converting enzyme inhibitor (ACEI) or an angiotensin II receptor antagonist (ARB), and a diuretic. Each medication is administered at maximum dosage or maximum tolerated dosage. If blood pressure remains above the target value or if ≥4 antihypertensive drugs are required in combination to achieve the target value, it is considered as RH [17].

### Statistical analysis

SPSS version 22 (SPSS Inc., Chicago, IL, USA) and R version 3.2.4 (R Foundation for Statistical Computing, Vienna, Austria) were used for the statistical analysis of . The Shapiro-Wilk test was used to determine whether the data were normally distributed. Results were expressed as mean±SEM or mean±SD for normally distributed data, and median ± interquartile range (25th to 75th percentiles) for nonnormally distributed data. The significance of differences between 2 groups was evaluated using the 2-tailed Student’s t test for normally distributed data or Mann-Whitney U test for nonnormally distributed data. The Fisher exact test or χ2 test was used to compare categorical variables. For all tests, a value of P<0.05 was considered to indicate statistical significance. A multivariate Cox’s proportional hazards model applying the adaptive least absolute shrinkage and selection operator (LASSO) was employed, and hazard ratios (HRs) were calculated using Cox regression analyses with 95% CIs or P-values. The ROC curve was used to evaluate the model using the area under the curve (AUC). its discriminatory ability was analyzed by determining the Harrell’s concordance index (C-index). The model’s calibration was represented by calibration plots. An individual risk score was derived from the nomogram for patients in the development cohort. The optimal cutoff point for each model was calculated for stratification of patients into low-risk and high-risk categories. The log-rank statistic was applied to determine the optimal cutoff point to provide the largest discrepancy in RH risk between the low-risk and high-risk groups. The Kaplan-Meier method was used to compare the incidence of RH between the two groups.

## Results

### Patient characteristics

A total of 1635 patients, who were diagnosed with HTN for the first time between 2019 and 2021 and achieved stable blood pressure after receiving medication treatment, were selected for this study. The data was then randomly divided into a Development cohort and a Validation cohort (Figure-1). The average age of the included population was 53 years old, with 70.08% being male and 34.01% currently smoking. The median resting blood pressure upon waking was 124/74mmHg. The median follow-up time was 31 months (15-36 months), and there were 373 cases (22.81%) of RH. The variables and event rates in the development cohort and validation cohort were similar, indicating good matching between the two groups (Table-1).

**Figure 1:**
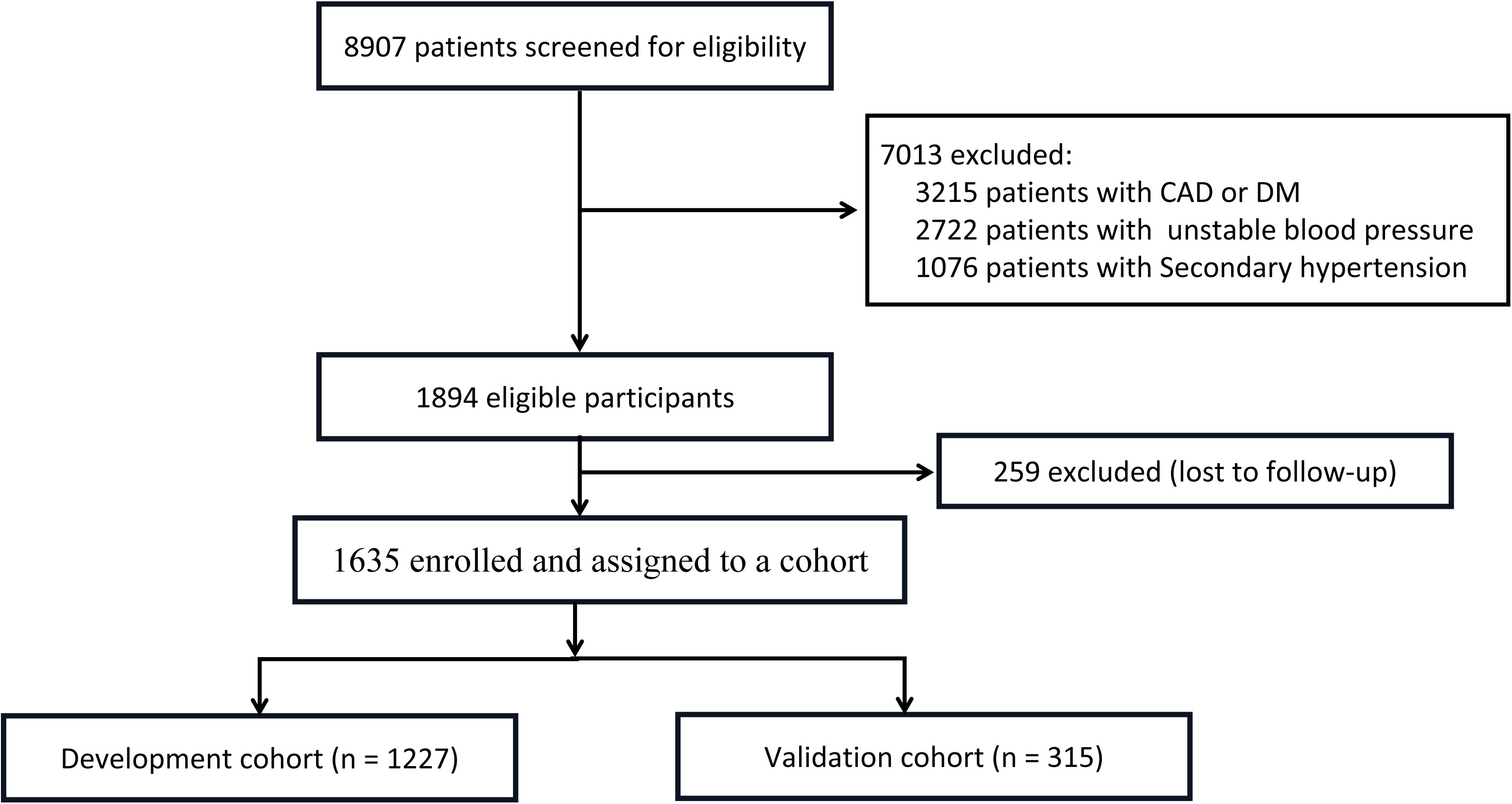
Inclusion of newly diagnosed primary hypertension and stable blood pressure after medication treatment Patients. Establishment of the Development Cohort (n = 1227) and the External Validation Cohort (n = 408)

**Table.1.** Baseline patient characteristics and Clinical outcome.

| Variable | All cohort<br>(N=1635) | Development cohort<br>(N=1227) | Validation cohort<br>(N= 408) | *P* |
| --- | --- | --- | --- | --- |
| **Characteristic** |  |  |  |  |
| Male sex, N % | 1146 (70.09) | 845 (68.87) | 301 (73.77) | 0.07 |
| Age, years | 53 ± 0.28 | 53 ± 0.33 | 54 ± 0.54 | 0.309 |
| Neck circumference, cm | 40.47 ± 0.11 | 40.4 ± 0.13 | 40.7 ± 0.23 | 0.83 |
| Body Mass Index, kg/m<sup>2</sup> | 28 ± 0.1 | 28 ± 0.12 | 28 ± 0.21 | 0.167 |
| Systolic blood pressure, mmHg | 124 ± 0.26 | 124 ± 0.3 | 125 ± 0.52 | 0.135 |
| Diastolic blood pressure, mmHg | 74 ± 0.2 | 74 ± 0.23 | 74 ± 0.45 | 0.083 |
| Current smoking, N % | 556 (34.01) | 414 (33.74) | 142 (34.8) | 0.695 |
| Current drinking, N % | 174 (10.64) | 130 (10.59) | 44 (10.78) | 0.914 |
| **Laboratory results** |  |  |  |  |
| Hemoglobin, g/L | 146 ± 0.39 | 145 ± 0.45 | 147.4 ± 0.77 | 0.267 |
| Blood platelet count, 10<sup>9</sup>/L | 232.49 ± 1.3 | 232.57 ± 1.41 | 231.13 ± 3.01 | 0.358 |
| Lymphocyte count, 10<sup>9</sup>/L | 2.06 (1.71, 2.47) | 2.06 (1.73, 2.45) | 2.07 (1.68, 2.51) | 0.833 |
| Neutrophil count, 10<sup>9</sup>/L | 4.89 (3.96, 6.46) | 4.91 (3.93, 6.41) | 4.86 (4.09, 6.77) | 0.128 |
| Alanine aminotransferase, U/L | 23.64 ± 0.25 | 23.89 ± 0.3 | 22.8 ± 0.41 | 0.625 |
| Aspartate aminotransferase, U/L | 22.69 ± 0.28 | 22.75 ± 0.34 | 22.51 ± 0.51 | 0.625 |
| Urea nitrogen, mmol/L | 5.01 ± 0.03 | 5.02 ± 0.03 | 4.99 ± 0.05 | 0.856 |
| Creatinine, µmol/L | 73 (64.2, 81) | 73 (64.5, 82) | 72 (64, 80) | 0.066 |
| Uric acid, µmol/L | 350.95 ± 1.98 | 352 ± 2.21 | 348 ± 4.4 | 0.253 |
| Total cholesterol, mmol/L | 4.3 (3.67, 4.97) | 4.3 (3.69, 4.93) | 4.28 (3.61, 5.1) | 0.791 |
| High-density lipoprotein cholesterol, mmol/L | 1.22 (1.08, 1.39) | 1.23 (1.08, 1.39) | 1.21 (1.07, 1.38) | 0.169 |
| Low-density lipoprotein cholesterol, mmol/L | 2.34 (1.92, 2.87) | 2.34 (1.92, 2.82) | 2.35 (1.91, 2.98) | 0.741 |
| Triglyceride, mmol/L | 1.55 (1.18, 2.13) | 1.6 (1.18, 2.13) | 1.43 (1.16, 2.04) | 0.083 |
| Fasting blood glucose, mmol/L | 5.54 ± 0.03 | 5.51 ± 0.03 | 5.62 ± 0.06 | 0.152 |
| Triglyceride glucose (TyG) index | 8.82 ± 0.01 | 8.82 ± 0.01 | 8.82 ± 0.03 | 0.457 |
| **Sleep monitoring** |  |  |  |  |
| Obstructive respiratory disturbance index | 15 (7.8, 28) | 15 (7.8, 26.4) | 15.2 (8.1, 30) | 0.415 |
| Average blood oxygen saturation | 92 (90, 93) | 92 (90, 94) | 92 (90, 93) | 0.146 |
| **End points** |  |  |  |  |
| Resistant_hypertension, N % | 373 ( 22.81) | 281 (22.9) | 92 (22.55) | 0.883 |

Data are presented as median (interquartile range), mean ± SD, or number (%).

### Potential predictors of resistant hypertension and development of a nomogram

In order to identify potential variables in the data, LASSO regression was performed on a set of 25 variables in the Development cohort. Non-zero parameter variables were selected, resulting in the identification of 8 variables with potential predictive value for RH (Figure-2, Figure-3). Further screening of these potential variables was conducted using multivariable COX regression analysis, leading to the final determination of 5 independent risk factors associated with developing RH: uric acid (HR=1.002, 95%CI: 1-1.002, P = 0.04), age (HR=1.001, 95%CI: 1-1.02, P = 0.04), AHI (HR=1.02, 95%CI: 1.01-1.02, P <0.01), fasting blood glucose (HR=1.28, 95% CI: 1.12-1.46, P <0.01), and TyG index (HR = 3.97, 95% CI: 2.81-5.62, P <0.01) (Figure-4). Subsequently, nomogram models were constructed based on these final five variables to predict the risk of developing RH at both month 31 and month 36 (Figure-5).

**Figure 2:**
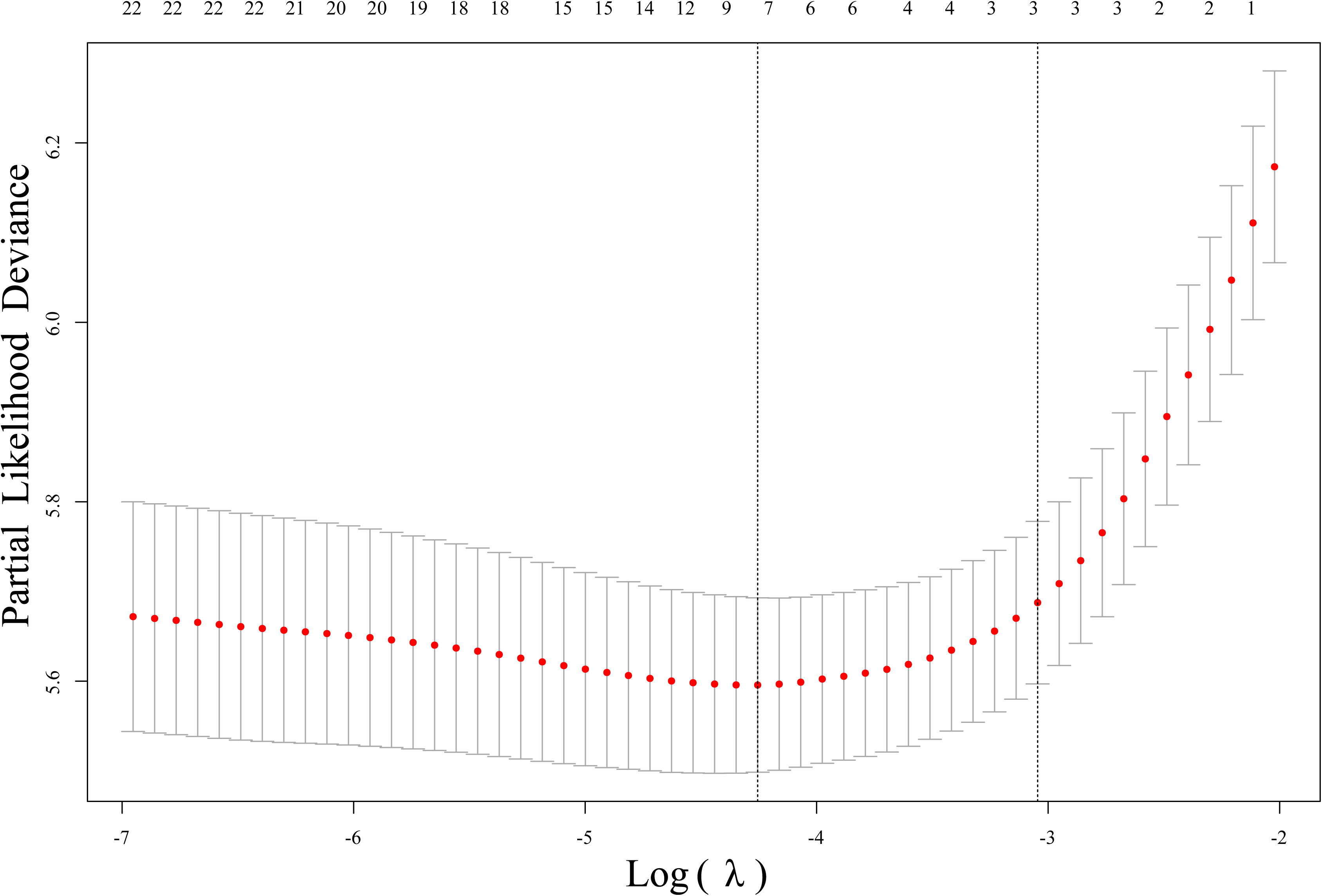
Showing the trends of 25 variables for long-term prognosis. The abscissa represents the optimal tuning parameter λ, and the ordinate represents the regression coefficient.

**Figure 3:**
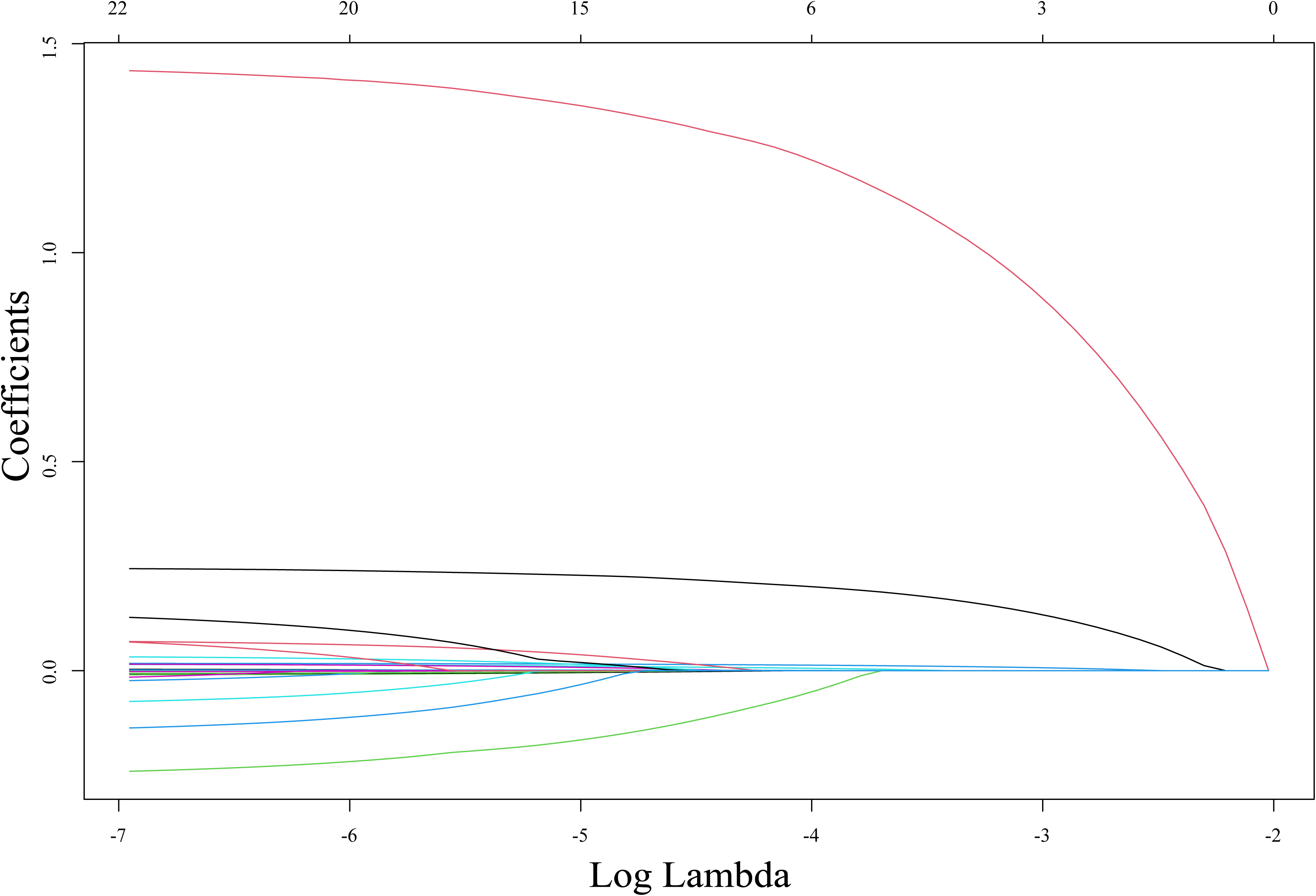
Illustrating the selection of the optimal tuning parameter λ and its relationship with the binomial deviation (binomial deviance). The vertical line indicates the optimized five nonzero coefficients derived by 10-fold cross-validation, leading to further multivariate Cox regression analysis.

**Figure 4:**
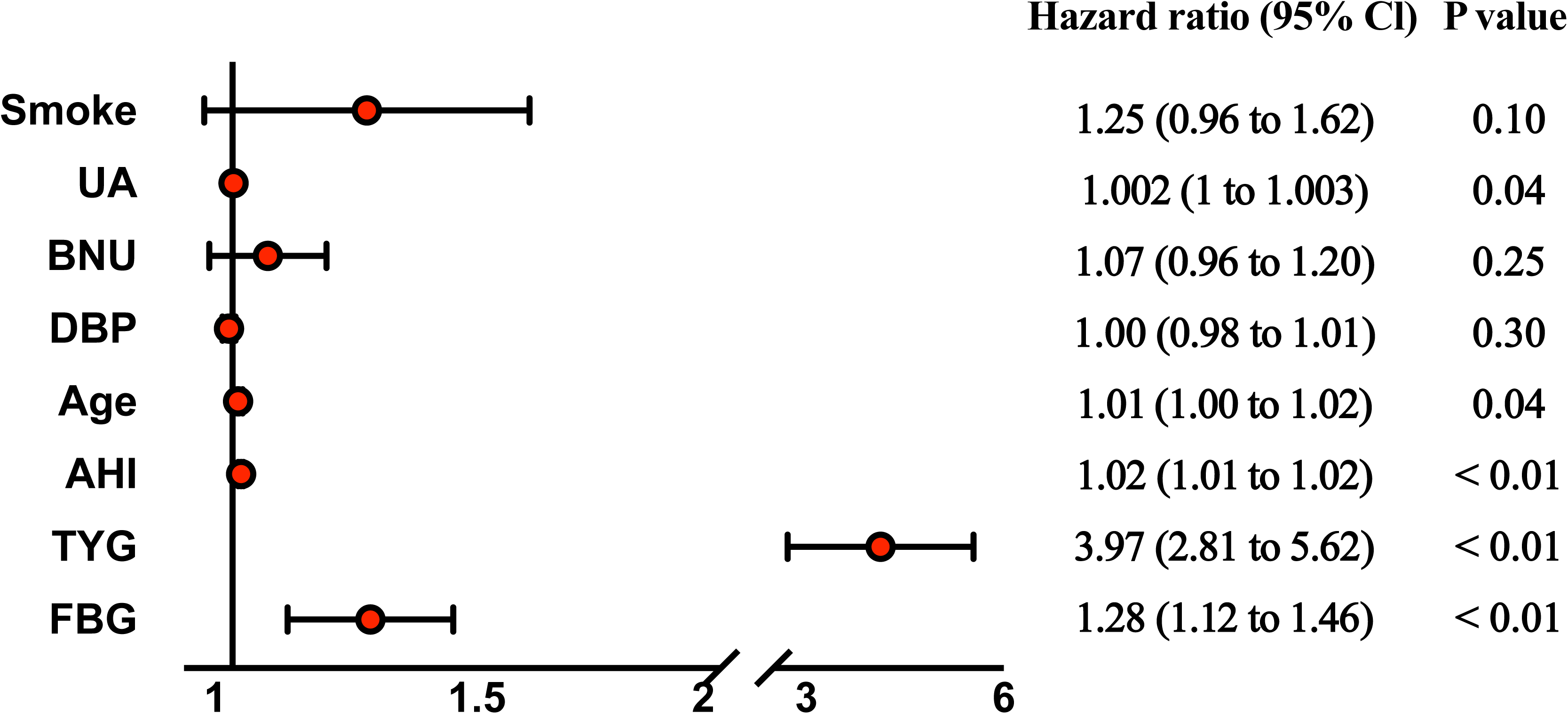
Displaying HRs for the independent prognostic variables identified by multivariate Cox regression in the development cohort.

**Figure 5:**
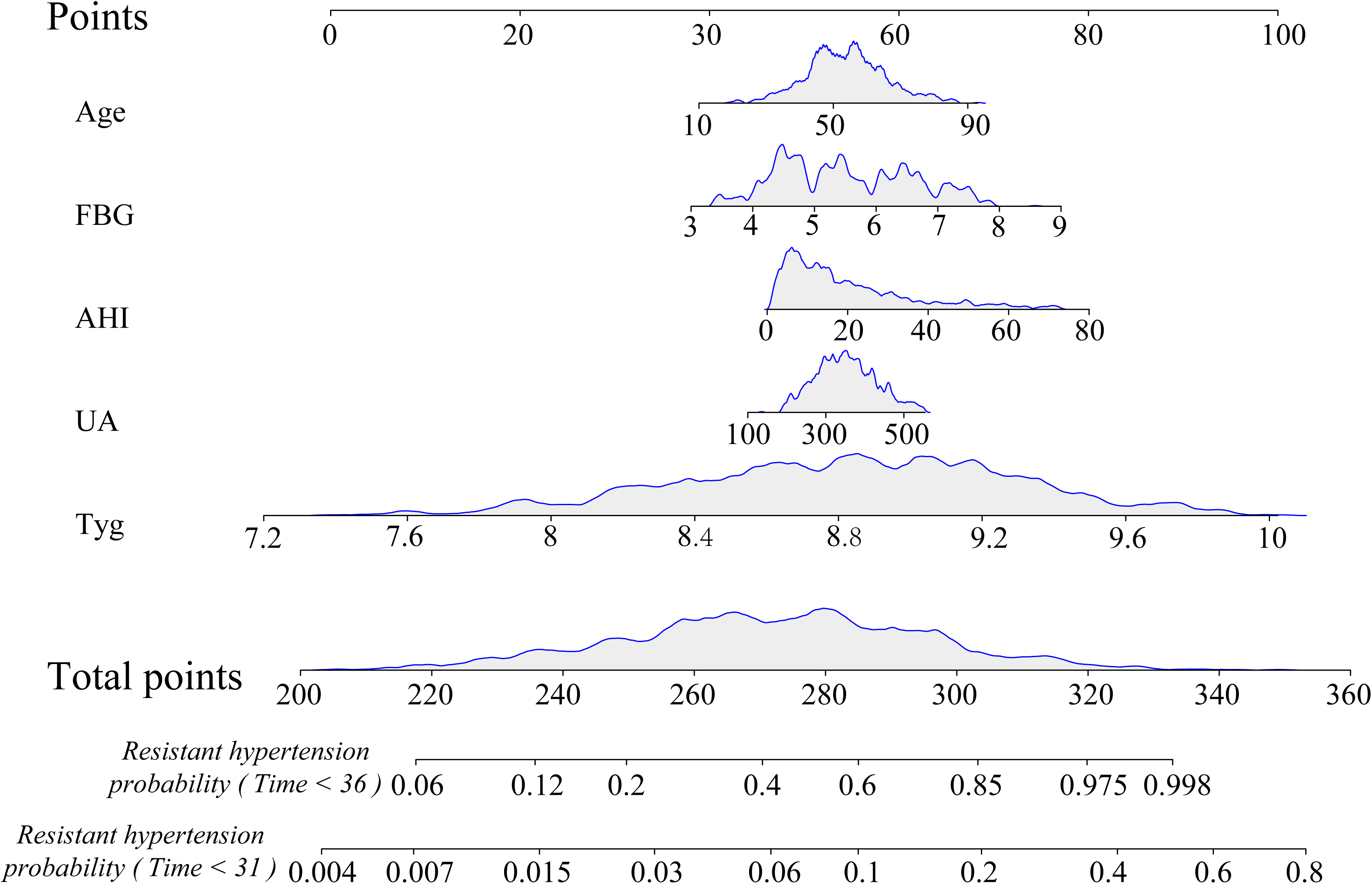
Demonstrating the nomogram used for predicting Resistant hypertension. The figure illustrates how to assign points to each patient based on their clinical characteristics, which are then summed into a total point score to calculate the probability of Resistant hypertension.

### Performance evaluation of the nomogram

To assess the predictive performance and stability of our model, we conducted a comprehensive analysis on the five selected variables in both the Development cohort and Validation cohort. Receiver Operating Characteristic (ROC) curves were generated to predict RH occurrence at 31 months and 36 months, with subsequent calculation of the area under the curve (AUC) for each time point (Figure-6). In the Development cohort, the AUC values were 0.895 and 0.837, respectively. Similarly, in the Validation cohort, the AUC values were 0.854 and 0.832, respectively. Subsequently, we employed calibration plots and calculated the C-index to assess the probability of developing RH. The C-index was found to be 0.76 in the Development cohort and 0.66 in the Validation cohort. Calibration plots exhibited concordance with both ROC curves and calibration curves (Figure-7). Finally, we quantified model risk for the overall population by utilizing median risk scores to distinguish between individuals at high-risk and low-risk levels. To compare occurrences of RH between these two groups, Kaplan-Meier analysis was conducted revealing that individuals classified as high-risk had a significantly higher likelihood of developing RH compared to those categorized as low-risk (Figure-8).

**Figure 6:**
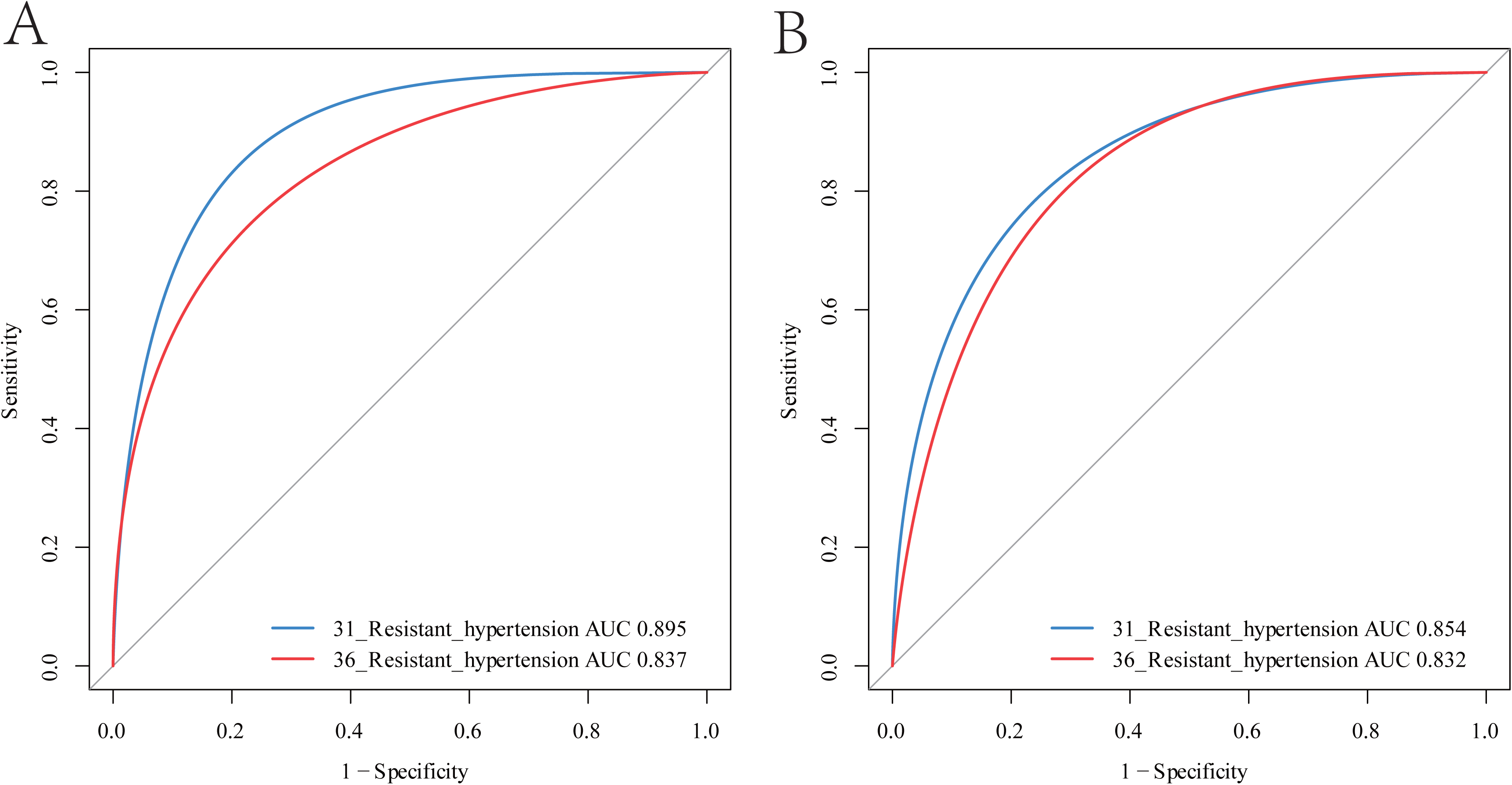
Evaluating the predictive accuracy of the nomogram for Resistant hypertension in both the development (A) and independent validation cohorts (B).

**Figure 7:**
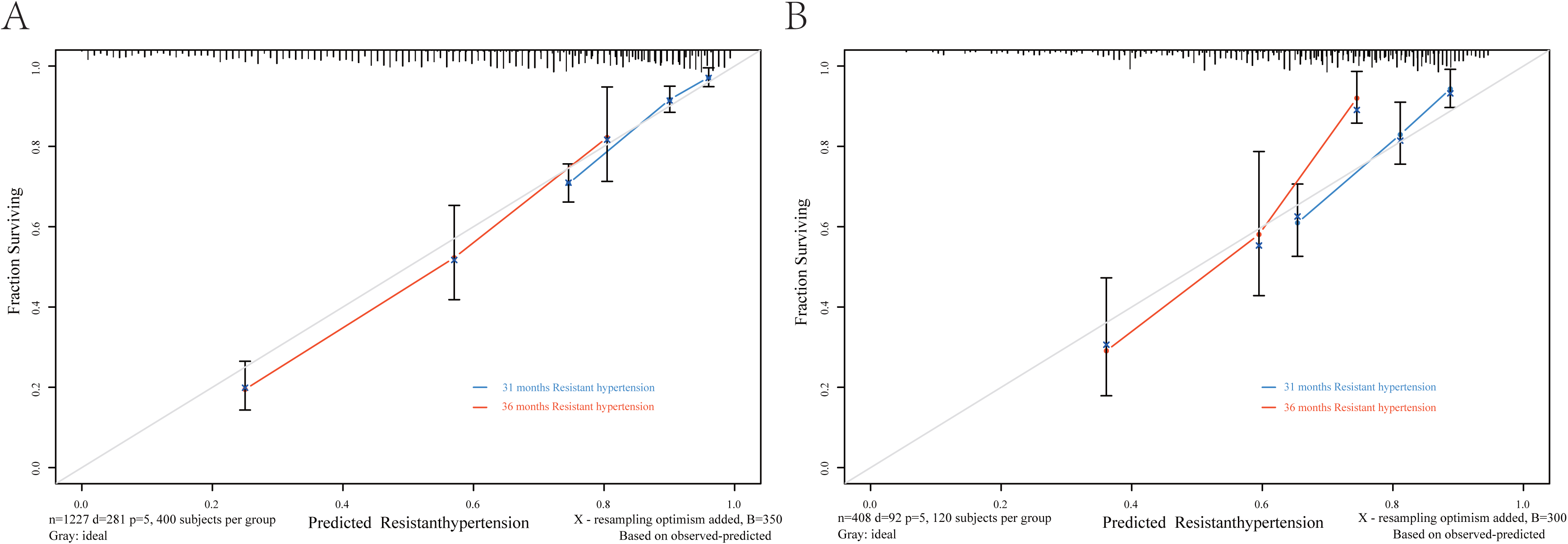
Depicting calibration curves for Resistant hypertension risk predictors in the development (A) and independent validation cohorts (B).

**Figure 8:**
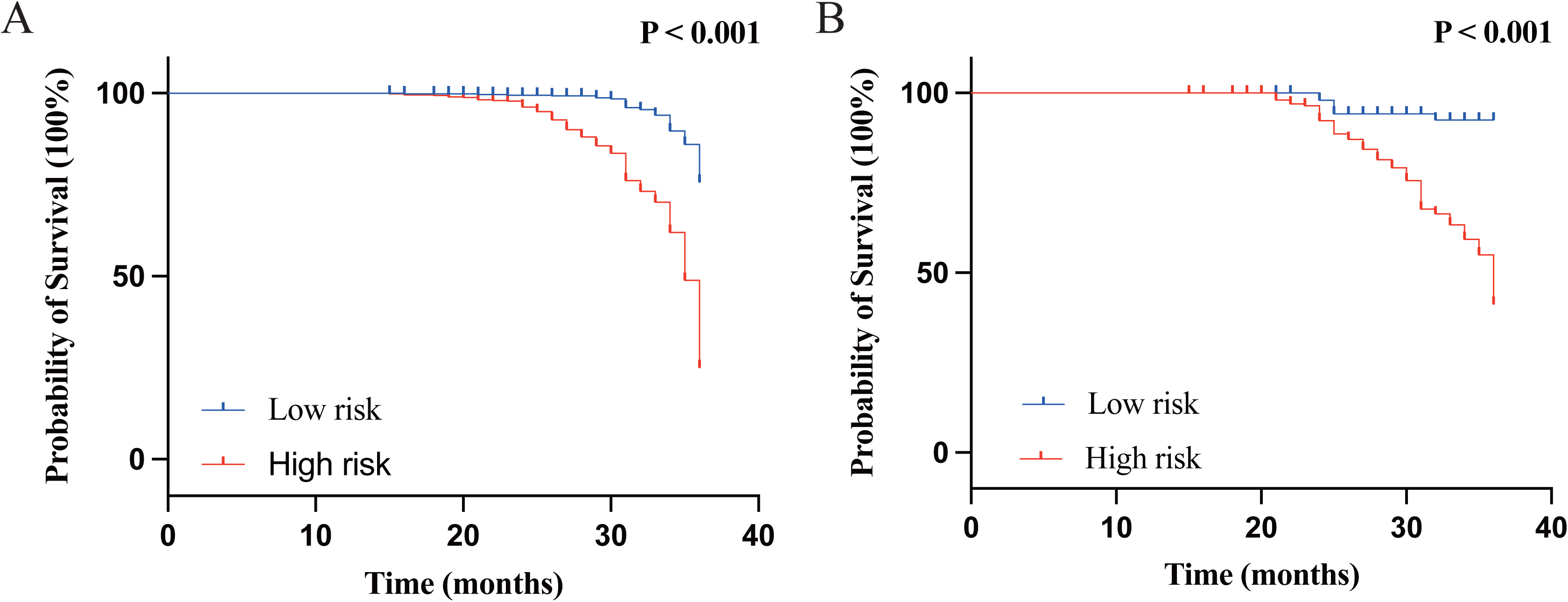
Illustrating the cumulative Resistant hypertension-free incidencel of patients in the development (A) and independent validation cohorts (B), stratified based on Resistant hypertension risk (high risk vs. low risk) using the median nomogram score.

## Discussion

Although significant progress has been made in the treatment of hypertension, there are still some patients who receive standard therapy but have poor blood pressure control. Currently, there is a lack of long-term prediction models for this subgroup of patients. In this study, we included patients with newly diagnosed HTN who achieved stable blood pressure after medication treatment and constructed a predictive model for RH through long-term follow-up. This is the first predictive model for RH that includes the TyG index along with several traditional risk factors (uric acid, age, AHI, and fasting blood glucose). According to the evaluation of model performance, the nomogram constructed in the development cohort was validated as effective in the validation cohort, indicating good predictive ability and stability of our model. This suggests that our model can provide risk stratification for patients with newly diagnosed primary hypertension and offer new insights into long-term antihypertensive treatment, which may support the development of new strategies.

The TyG index can serve as an indicator of IR in the body [18,19]. It has been identified as an important diagnostic marker for predicting metabolic syndrome [20]. The formation and development of RH is a dynamic process. In this study, we have confirmed for the first time that the TyG index is the most significant risk factor for RH, while also providing evidence that IR may contribute to its occurrence and progression. Current research suggests that the main causes of RH are increased blood volume, enhanced sympathetic nervous system activity, and the influence of cortisol and pro-inflammatory factors [21–23]. Understanding how these pathological changes occur in patients with RH is crucial for its treatment. IR may play a key role among these factors. It is well known that renal epithelial cells are sensitive to insulin and that insulin has an antinatriuretic effect. IR primarily promotes sodium-potassium reabsorption in distal renal tubules by directly or indirectly increasing renin-angiotensin-aldosterone system activity, leading to increased reabsorption of sodium, potassium, and water in renal tubules and resulting in increased blood volume and cardiac output [24,25]. IR can also disrupt sympathetic nervous system function, increase peripheral and renal vascular resistance, impair endothelial function, cause lipid disorders and atherosclerosis; all of which are critical factors influencing RH [26,27]. Existing clinical studies have also found that IR is an independent risk factor for autonomic imbalance based on 6-month follow-up observations [28]. Furthermore, research has shown a close relationship between IR and cortisol levels as well as pro-inflammatory factors; suggesting involvement of hypothalamic-pituitary-adrenal axis activity and macrophages in fat mobilization [29–30]. Therefore,this may be why the TyG index outperforms other risk factors.

In addition, our study also found that age, AHI (Apnea-Hypopnea Index), uric acid levels, and fasting blood glucose are independent risk factors for RH. AHI is an evaluation index of the severity of respiratory sleep apnea. There have been numerous reports on the association between sleep apnea and RH. However, it is worth noting that about half of primary HTN patients have obstructive sleep apnea, while about half of obstructive sleep apnea patients have primary HTN; these two conditions often coexist [31,32]. Uric acid is the main endogenous antioxidant in the human body as a final product of purine metabolism. However, impaired synthesis and excretion of uric acid can lead to hyperuricemia, which in turn induces endothelial dysfunction and contributes to the development of RH [33]. Studies have also confirmed a close correlation between fasting blood glucose levels and blood pressure levels and identified it as an independent risk factor for elevated blood pressure [34]. Furthermore, we found that patient age, AHI score, uric acid levels, and fasting blood glucose levels all influence IR [35–39]. This may explain why IR has the highest HR value among quantifiable indicators in independent risk factors while also suggesting that IR is a major cause for the progression from high blood pressure to RH.

In our study, a total of 1635 patients were included in the overall population, all of whom were newly diagnosed with HTN and achieved stable blood pressure through medication treatment. During the recruitment phase, it was found that patients with respiratory sleep apnea refused to use non-invasive ventilator therapy, making it impossible to differentiate between primary and secondary hypertension. In order to reduce sample bias, we excluded patients who agreed to use non-invasive ventilator therapy. Additionally, since diabetes and coronary heart disease have a promoting effect on the progression of HTN and there is antagonism among the drugs used, we excluded patients with diabetes and coronary heart disease throughout the entire study in order to solely investigate the process of HTN progressing into RH.

There are certain limitations in this study. Firstly, the sample size is small and the research data are all from the Fifth Affiliated Hospital of Xinjiang Medical University. Although internal validation was conducted in this study, a large-scale multicenter prospective cohort study is still needed for verification. Secondly, the diagnosis of RH requires adequate adjustment of medication, and individualized treatment plans may affect the results. Thirdly, while the predicted results in the survival curve remained unchanged over time in this study, in reality, disease outcomes may vary due to improvements in treatment, early detection, and changes in natural course of illness. Therefore, as time goes on, the performance of survival curves may become inaccurate.

### Conclusions

This study developed a prognostic model for patients with difficult-to-treat HTN after stable blood pressure was achieved through initial diagnosis and medication treatment. The model combines the TyG index with recognized clinical risk factors. Our data validated that the TyG index can serve as a quantitative long-term risk biomarker. The developed predictive nomogram can be easily applied in clinical practice, supporting individualized management of hypertensive patients.

## Funding

This research was funded by the National Natural Science Foundation of China (grant number 82170345).

## Author Contribution

YY.Z and X.X conceived and conducted the study. HT.Y and JK.L conducted the clinical study and authored the manuscript. HT.Y and Y.Y were responsible for data collection and analysis. All authors contributed to the article and approved the final version. HT.Y and JK.L made equal contributions to this work.

## Conflict of Interest

The authors declare that they have no conflict of interests.

## Ethics approval and consent to participate

The study protocol was approved by the Ethics Committee of the Fifth Affiliated Hospital of Xinjiang Medical University (2022XE0103-1). Due to the retrospective design of the study, the requirement for obtaining informed consent from eligible patients was waived by the ethics committee.

## Data sharing agreement

Individual participant data that support the findings of this article, following the process of de-identification, are available from the corresponding author upon reasonable request

## Notes

### Competing Interest Statement

The authors have declared no competing interest.

### Clinical Trial

ChiCTR1900025479

### Author Declarations

The study protocol was approved by the Ethics Committee of the Fifth Affiliated Hospital of Xinjiang Medical University (2022XE0103-1).

